# Nicotine pouch adverts reach 12 times more young men than women: targeting and reach on Meta social media platforms in the UK

**DOI:** 10.64898/2026.05.27.26354221

**Authors:** Haoxiong Sun, Sarah E. Jackson, Leon Y. Xiao, Sharon Cox, Melissa Oldham, Jiang Yu, Harry Tattan-Birch

## Abstract

**Aims:** To examine which demographic groups nicotine pouch advertisers chose to target on social media, and which groups Meta’s algorithms actually delivered the adverts to.

**Design:** Cross-sectional analysis of advert-level data from the Meta Ad Library.

**Setting:** Meta social media platforms (including Facebook and Instagram) in the UK.

**Cases:** A sample of 642 nicotine pouch adverts identified by keyword search, shown in the 12 months up to December 2025, and a comparison sample of 1,125 general adverts. Analyses of reach were restricted to adverts with no advertiser-set gender restriction that targeted all adult ages (373 pouch adverts; 674 general).

**Measurements:** Outcomes, both extracted from the Meta Ad Library, were advertiser-set gender and age-group targeting criteria (i.e., groups eligible to be shown each advert) and estimated advert reach to each group (i.e., number of people who saw each advert). Male-to-female reach ratios (RR) within age groups, and reach ratios comparing age groups, were calculated per advert and summarised using geometric means. To assess whether patterns were pouch-specific, comparisons with general adverts were made using ratios of reach ratios (RRR).

**Findings:** Advertisers of nicotine pouches targeted a broad sample. Most adverts 81.0% (520/642) had no gender restriction, the remainder were restricted to men only. All were restricted to adults (minimum age 18 years) and most (96.9%; 622/642) had no upper age limit. Despite this, of pouch adverts eligible to be shown to all adults, adverts were more likely to reach men, particularly among younger men. Among 18–24-year-olds, pouch adverts reached around twelve times as many men as women (RR 12.5, 95% CI 10.9–14.4), compared with a slight skew towards women for general adverts (RR 0.81, 95% CI 0.70–0.95), corresponding to an RRR of 15.4 (95% CI 12.3–18.8). Pouch adverts also showed a skew in reach towards younger age groups. Relative to those aged 35–44 years, reach was higher among 18–24-year-olds for nicotine pouch adverts (RR 1.50, 95% CI 1.31–1.71) but much lower for general adverts (RR 0.19, 95% CI 0.17–0.21), corresponding to an RRR of 7.9 (95% CI 6.6–9.5).

**Conclusions:** Nicotine pouch adverts on social media were often eligible to be shown broadly to all demographic groups but were disproportionately delivered to young men.

## Introduction

Oral nicotine pouches are small, tobacco-free products placed between the gum and lip that deliver nicotine without containing tobacco leaf, and come in a wide range of flavours, strengths, and packaging styles (1). The use and sales of these particular pouches have risen sharply in recent years (2–4). They expose users to fewer toxicants than cigarettes, but they are not risk-free. Their public health impact will depend on who uses them, trajectories of use and their long-term impact on health outcomes (2, 5). Use of nicotine pouches in Great Britain has increased since 2020, with the steepest rises among young people, especially young men (4). By 2025, 7.5% of 16–24-year-old men reported current pouch use, compared with 1.9% of women the same age and 1.0% of adults overall (4), but it is not yet clear what is driving these sociodemographic differences in use rates. One possibility is that how nicotine pouch advertising reaches different groups may contribute to use being concentrated in young men.

Unlike cigarettes, and to a lesser extent vapes, nicotine pouches have sat in a relatively lightly regulated space in the United Kingdom (UK) (4–6). Restrictions on their advertising have been limited, and marketing has become widespread across out-of-home advertising, sponsorships, and social media (6, 7). At the time of the study, nicotine pouches in the UK had no statutory minimum age of sale and were subject only to general advertising codes (8, 9). The Tobacco and Vapes Act 2026 will introduce an age of sale of 18 and an advertising ban (10). Social media advertising differs from traditional advertising: platforms do not simply show adverts to the groups selected by advertisers but use optimisation systems (e.g. an algorithm) that influence which groups adverts actually reach (11, 12). These systems learn from user engagement like clicks or dwell time, to deliver adverts to people predicted to respond, which can shift reach away from advertiser-set criteria. As a result, adverts set to be shown broadly may in practice reach some demographic groups much more than others. These reach patterns are determined by platform algorithms, which are not observable to advertisers, regulators, or the public.

This distinction between targeting and reach is important for public health and regulation. Current rules often focus on advertiser-set targeting criteria, such as minimum age, but these settings might not reflect which groups are actually reached once adverts are distributed on the platform (10, 11). For example, Dutch gambling advertising regulations prohibit social media adverts from targeting young adults aged 18–23 years (13). If regulation is based only on advertiser inputs, it may be ineffective, missing an important part of how exposure is distributed.

The Meta Ad Library provides an opportunity to examine both sides of this process: it documents the age and gender groups advertisers selected as eligible to be shown an advert, and the demographic breakdown of estimated reach among people who saw it (12, 14). Therefore, in this study we aimed to examine which groups nicotine pouch adverts were targeted to and which groups they reached on Meta platforms in UK in 2024-2025. Specifically, our objectives were to:

1. Describe which age and gender groups advertisers targeted as eligible to be shown nicotine pouch adverts.
2. Assess whether nicotine pouch adverts with no gender restriction were more likely to reach men than women, and whether this differed from general adverts.
3. Assess whether nicotine pouch adverts eligible to be shown to all adults were more likely to reach younger than older age groups, and whether this differed from general adverts.

## Method

### Data source

We used data from the Meta Ad Library, a publicly accessible database of adverts run on Meta platforms. For adverts delivered to the UK and European Union, the Meta Ad Library API provides estimated information on ad delivery, including targeting parameters and demographic breakdowns of reached audiences by age and gender (14). Throughout this paper, we use ’United Kingdom (UK)’ to refer to data and policies covering England, Scotland, Wales, and Northern Ireland, and ’Great Britain (GB)’ to refer to data covering England, Scotland, and Wales only.

We retrieved data on adverts shown in the UK on Meta platforms during the 12 months preceding 8 December 2025. This 12-month window represents the most recent complete year of advertising at the time of extraction, covering a full annual cycle, and corresponds approximately to the period for which non-political adverts remain available in the Ad Library. Advertisements potentially related to nicotine pouches were identified by searching the Ad Library API with the search term "nicotine pouch", using Meta’s unordered keyword matching. This term was chosen because it directly names the product category, and adverts returned by the search but not related to nicotine pouches were excluded following manual review.

In addition, a comparison sample of general adverts, unrelated to nicotine pouches, was identified over the same period. A pool of advertiser pages was first assembled by searching the Meta Ad Library using six broad, non-topical seed terms (’a’, ’e’, ’the’, ’and’, ’shop’, ’new’) as a proxy for random sampling, since the API does not support retrieval of random advertiser pages directly. This yielded a pool of 221 unique advertiser pages.

From this pool, 150 pages were randomly sampled. Advertisements delivered by the sampled pages during the same 12-month window were retrieved via the API in batches of up to 10 pages, with retrieval capped at 75 adverts per batch. Of the 150 pages, 36 returned adverts, yielding 1,125 general adverts. For each advert, we extracted advertiser-set targeting parameters (including age, gender, and location), along with demographic breakdowns of estimated reach.

### Sample

The search returned 923 potential nicotine pouch advertisements. Each advert was classified as promoting nicotine pouches or not by one author (HS). A second author (YJ) independently coded a random sample of 80 adverts, blind to the first coder’s classifications. Agreement was 92.5% (74 of 80 adverts; Cohen’s κ = 0.81), and disagreements were resolved by discussion. Both coders classified adverts on the basis of advert content only, without reference to targeting or reach data.

Classification was made at the level of the advertiser page, using the advert text, link titles and descriptions, and web checks of brand identity where needed, and adverts promoting nicotine-containing pouches by manufacturers or retailers, or as sponsored editorial content, were included. Adverts whose content had been removed by Meta were classified on the basis of other adverts from the same page, and excluded where no such evidence existed.

In total, 281 adverts were excluded, leaving 642 adverts for analyses of targeting. Analyses of reach focused on adverts with no advertiser-set gender restriction that targeted all adult ages (i.e., a minimum age of 18 with no upper limit) and reported reach for men and women in the UK (n = 373), so that any demographic skew in reach reflects platform delivery rather than advertiser targeting.

A sample of 1,125 general adverts was also identified. Of these, 682 had no advertiser-set gender restriction and targeted all adult ages, and 8 were excluded because no UK-specific reach breakdown was provided for them, leaving 674 used as the comparison group. The details of inclusion and exclusion procedures of both samples are shown in **Supplementary Figure 1**.

### Measures

Advertiser-set targeting measures included the minimum and maximum age and the genders selected as eligible to be shown each advert. Meta permits adverts to be shown to users aged 13 and over, with only age and location available as targeting criteria for 13–17-year-olds (15). Gender in the Ad Library reflects the gender recorded on users’ accounts and is reported by Meta as male, female or unknown.

Reach of each advert was available by age group (18-24, 25-34, 35-44, 45-54, 55-64, 65+) and gender (male and female users and a small ’unknown’ category, which accounted for about 1% of reach in both samples and was not analysed). No reach was reported for 13-17-year-olds for any advert, because every advert had an advertiser-set minimum age of 18 years or above. Reach was measured using Meta’s estimated reach metrics, which count the number of unique people who saw each advert at least once. Meta deduplicates reach across linked Facebook and Instagram accounts at the user level, broken down by age group and gender (14).

Where an advert had zero reported reach for one gender within a specific age bracket, or zero reach within a specific age group, a continuity correction of 0.5 was added before computing the relevant ratio (Haldane-Anscombe correction), so that adverts with zero counts were retained (16).

### Analysis

Analyses were conducted at the level of the advert. This approach estimates the typical reach pattern of an advert, rather than being dominated by a small number of adverts with very high reach.

We summarised advertiser-set targeting by describing the distribution of minimum and maximum ages selected for each advert, and whether adverts had no gender restriction or were restricted to men or women only.

Among adverts with no gender restriction, we compared how often adverts reached men and women within each age group. For each advert, we calculated the ratio of reach (RR) to men versus women and summarised these using geometric means.

To assess whether this pattern differed from general advertising on the platform, we repeated the same analysis for general adverts and compared results by calculating the ratio of reach ratios (RRR). This comparison isolates pouch-specific reach patterns from baseline platform demographics, and RRR greater than 1 indicates a stronger male skew in nicotine pouch reach relative to general advert reach.

We then performed an analogous analysis of how reach varied across age groups. For each advert, reach was summed across genders within each age group, and we calculated the ratio of reach in each age group to reach in those aged 35-44, after applying the same continuity correction (adding 0.5 to both the age-group and reference-group reach counts). These ratios were summarised using geometric means. We compared these patterns with general adverts by calculating the RRR.

Bootstrap resampling of adverts (1,000 iterations) was used to construct 95% confidence intervals, calculated from the 2.5th and 97.5th percentiles (17). As a sensitivity analysis, we repeated the bootstrap by resampling advertiser pages rather than individual adverts. All adverts from each selected page were included together in each resample, to account for clustering of adverts within advertisers. We also report male-to-female ratios based on total reach summed across adverts, which give greater weight to adverts with higher reach (Supplementary Table 4). Data extraction and sampling were conducted using Python (version 3.13) interfacing with the Meta Graph API (v24.0). Statistical analyses were conducted in R (version 4.5.0). The analyses were not pre-registered and should be considered exploratory.

## Results

A total of 642 nicotine pouch-related adverts and 1,125 general adverts were identified that ran across Meta platforms in the UK in the year up to 8 December 2025. Pouch-related adverts were run by 70 unique advertiser pages, and the general adverts were retrieved from 36 advertisers (**Supplementary Table 1**). The composition of the general advert sample by advertiser category is shown in **Supplementary Table 2**.

### Targeting

Table 1 summarises the age and gender groups advertisers selected as eligible to be shown each advert. Most nicotine pouch adverts (81.0%; n=520) had no advertiser-set gender restriction and were eligible to be shown to users of any gender. The remaining 122 adverts (19.0%) were restricted by the advertiser to men only, and none were restricted to women only. All nicotine pouch adverts were restricted to adults (≥18 years). Most (93.8%; n=602) set the minimum age at 18 years, while a small number (6.2%; n=40) set a higher minimum age of 25 years or above. Almost all adverts (96.9%; n=622) had no upper age limit.

**Table 1.** Advertiser-set targeting criteria specifying which genders and age groups were eligible to be shown nicotine pouch adverts on Meta platforms (N=642).

| **Targeting criterion** | **Category** | **n of adverts** | **% of adverts** |
|---|---|---|---|
| Gender | No gender restriction | 520 | 81.0 |
|  | Male only | 122 | 19.0 |
|  | Female only | 0 | 0.0 |
| Minimum age | No lower limit | 0 | 0.0 |
|  | Minimum age 18 years | 602 | 93.8 |
|  | Minimum age ≥25 years | 40 | 6.2 |
| Maximum age | No upper limit | 622 | 96.9 |
|  | Under 65 years | 3 | 0.5 |
|  | Under 55 years | 15 | 2.3 |
|  | Under 45 years | 2 | 0.3 |

Note: Targeting refers to the criteria set by advertisers in Meta’s ad system, specifying which users were eligible to be shown an advert. It does not indicate which groups actually saw the advert. All adverts specified a minimum target age of at least 18 years. Adverts included were those shown in the UK on Meta platforms in the 12 months preceding 8 December 2025.

### Reach by gender

Of the 642 pouch adverts, 373 had no advertiser-set gender restriction, targeted all adult ages, and were used in analyses of reach. Among these 373 advertisements, estimated reach was greater for men than for women across all age groups (**Figure 1**, **Supplementary Table 3, 4**). This difference was largest at younger ages: in the 18–24 age group, adverts reached around twelve times as many men as women on average (RR 12.5, 95% CI 10.9 to 14.4), whereas the difference was smaller among those aged 65 years and over, where adverts reached around three times as many men as women (RR 3.0, 95% CI 2.7 to 3.5).

**Figure 1.**
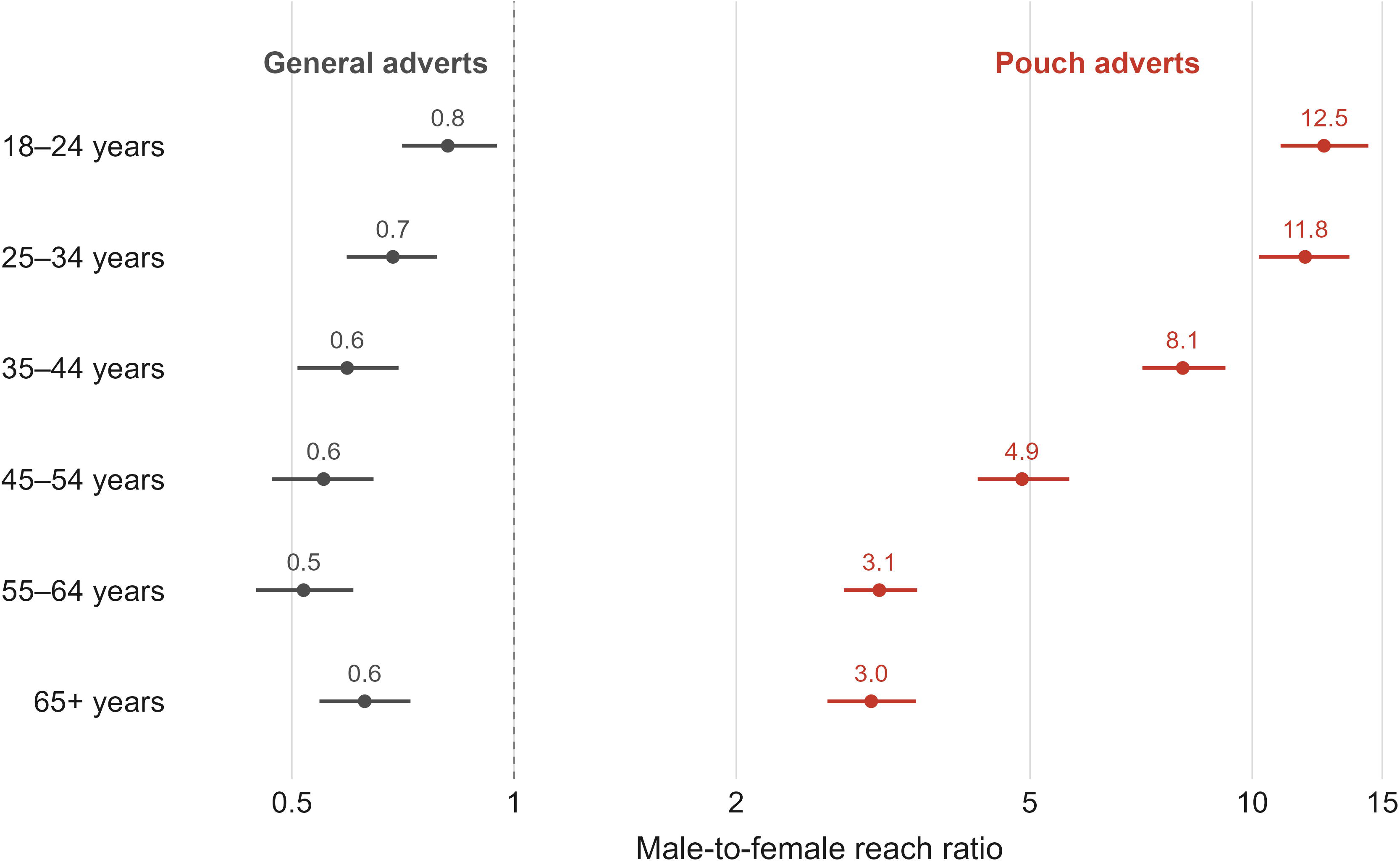
Male-to-female reach ratios (RR) by age group for nicotine pouch adverts (n=373) versus general adverts (n=674) on Meta platforms. Values >1 indicate greater reach to men. Estimates are geometric means across adverts. Horizontal bars indicate 95% confidence intervals from bootstrap resampling. Analyses restricted to adverts with no advertiser-set gender restriction that targeted all adult ages.

In contrast, general adverts unrelated to nicotine pouches that had no advertiser-set gender restriction and targeted all adult ages (n=674) reached similar numbers of men and women among 18–24-year-olds and fewer men than women at older ages (**Figure 1**, **Supplementary Table 3**). Taking this underlying platform pattern into account, nicotine pouch adverts showed a stronger skew in reach towards men across all age groups. In the 18–24 age group, the male-to-female relative difference in reach was around fifteen times larger for nicotine pouch adverts than for general adverts (RRR 15.4, 95% CI 12.3 to 18.8). A similar pattern was seen in the 25–34 (RRR 17.2, 95% CI 14.0 to 21.2) and 35–44-year-old (RRR 13.6, 95% CI 11.0 to 16.6) age groups. The relative difference was smaller at older ages but remained clear, with the male-to-female difference in reach still around five times larger among those aged 65 years and over (RRR 4.9, 95% CI 4.0 to 5.9).

Median estimated total reach per advert was 614 (interquartile range [IQR] 82–7,464; range 1–2,118,402) for nicotine pouch adverts and 515 (IQR 119–1,777; range 1–418,312) for general adverts. Despite this variation, a similar male skew was observed when reach ratios were instead calculated from total reach summed across all adverts, which gives greater weight to adverts with higher reach (**Supplementary Table 4**).

Results were similar when the bootstrap resampled advertiser pages (**Supplementary Table 5**).

### Reach by age

Nicotine pouch adverts were also more likely to reach younger than older age groups (**Table 2**). Compared with those aged 35–44 years, adverts were around 1.5 times more likely to reach 18–24-year-olds (RR 1.50, 95% CI 1.31 to 1.71) and around twice as likely to reach 25–34-year-olds (RR 2.02, 95% CI 1.87 to 2.18). In contrast, adverts were less likely to reach older age groups, with relative reach around half as high among those aged 45–54 years (RR 0.54, 95% CI 0.50 to 0.58) and lower still among older groups.

**Table 2.** Age-group reach ratios (RR) (vs 35-44) for pouch adverts and general adverts, and the ratio of reach ratios (RRR)

| Age group | RR |  | RRR |
|---|---|---|---|
|  | Pouch adverts (95% CI) | General adverts (95% CI) | Pouch vs general (95% CI) |
| 18–24 | 1.50 (1.31, 1.71) | 0.19 (0.17, 0.21) | 7.91 (6.56, 9.50) |
| 25–34 | 2.02 (1.87, 2.18) | 0.82 (0.76, 0.88) | 2.46 (2.20, 2.75) |
| 35–44 | Ref | Ref | Ref |
| 45–54 | 0.54 (0.50, 0.58) | 0.81 (0.74, 0.87) | 0.67 (0.59, 0.75) |
| 55–64 | 0.31 (0.27, 0.35) | 0.67 (0.60, 0.76) | 0.46 (0.38, 0.56) |
| 65+ | 0.26 (0.22, 0.30) | 0.39 (0.34, 0.46) | 0.66 (0.53, 0.82) |

**Note:** RR = reach ratio, RRR = ratio of reach ratios. RRs compare reach of adverts to other age groups versus 35-44 (values >1 indicate greater reach than the 35–44 reference group). RRRs compare this difference for nicotine pouch adverts versus general adverts.
Analyses were restricted to adverts with no advertiser-set gender restriction that targeted all adult ages. Estimates are geometric means across adverts with 95% confidence intervals (CIs) from bootstrap resampling. N for pouch adverts = 373, and N for general adverts = 674.

Among general adverts unrelated to nicotine pouches, a different pattern was observed. Relative to those aged 35–44 years, adverts were much less likely to reach younger age groups and slightly less likely to reach older age groups (**Table 2**).

When accounting for these patterns, nicotine pouch adverts showed a much stronger skew in reach towards younger age groups than general adverts. In the 18–24-year-old group, the relative difference in reach compared with 35–44-year-olds was around eight times larger for nicotine pouch adverts than for general adverts (RRR 7.91, 95% CI 6.56 to 9.50). A smaller but still clear difference was seen in the 25–34-year-old group (RRR 2.46, 95% CI 2.20 to 2.75). In contrast, nicotine pouch adverts showed lower relative reach across all groups aged 45 years and over (RRR <1).

When accounting for clustering of adverts within advertisers, the skew towards younger age groups remained evident (RRR for 18–24 vs 35–44 years: 7.91, 95% CI 3.96 to 15.71; 25–34 vs 35–44 years: 2.46, 95% CI 1.38 to 4.18), but for groups aged 45 and over the intervals were wide and compatible with both weaker and stronger age skews than among general adverts **(Supplementary Table 6).**

## Discussion

In this study, we examined the targeting and reach of nicotine pouch advertisements, and two distinct patterns emerged. First, one in five adverts was explicitly restricted by advertisers to men, reflecting deliberate targeting choices. Second, adverts that advertisers had set to be shown to all adults were nonetheless delivered far more often to men, particularly younger men: in the 18–24 age group, pouch adverts reached around twelve times as many men as women, far exceeding the male-to-female reach ratio for general adverts on the same platform. This second pattern reflects platform delivery rather than advertiser instruction, and is particularly striking given that women outnumber men on Facebook and Instagram and spend more time on these platforms, so that advertising on Meta would, in the absence of algorithmic skew, be expected to reach more women than men (11). Reach was also concentrated among younger age groups, with the skew towards 18-24-year-olds approximately eight times larger for nicotine pouch adverts than for general adverts.

These findings are consistent with broader evidence that algorithmic ad delivery systems do not simply reflect advertiser-set targeting parameters but actively shape which groups see an advert (11, 18). In the case of nicotine pouches, our findings are consistent with preferential delivery to users predicted by the platform to be more likely to engage with this content, concentrating exposure among young men without explicit instruction from advertisers (11, 18). Other explanations are also possible.

Advertisers may have used imagery and themes that appealed particularly to men, pouch brands’ existing followers may have been predominantly male, or interest-based targeting not reported in the Ad Library may itself have been male-skewed. Future research linking advert content and other targeting inputs to audience reach could help distinguish these explanations. Similar discrepancies have been found in other product categories, including video game advertising, where adverts set to reach broad audiences were disproportionately delivered to specific demographic groups (12, 19).

The demographic pattern of reach we observed closely mirrors the pattern of nicotine pouch use in Great Britain, where prevalence has risen most sharply among young men (4). This raises the possibility that advertising exposure and use may be mutually reinforcing. However, we cannot establish the direction of this relationship from these data alone: it is possible that the algorithm targets young men because they engage more with pouch content, that advertising exposure contributes to higher use among young men, or both.

This issue is particularly relevant in the UK at present. Nicotine pouches have fallen outside the statutory advertising restrictions that apply to tobacco products, with oversight focused on advertiser-set targeting parameters rather than on the demographic groups adverts are actually delivered to (8, 9). The UK’s Tobacco and Vapes Act, which received Royal Assent and passed into law in April 2026, is intended to bring nicotine pouches within restrictions on advertising, promotion, and sponsorship that apply to other nicotine and tobacco products (20). However, even under such a framework, regulation focused solely on targeting inputs may not capture how exposure is distributed across demographic groups (11, 21). Minimum ages of sale and advertising rules for nicotine pouches vary internationally, so advertiser-set targeting patterns may differ in other jurisdictions, although similar platform delivery mechanisms may operate across markets (22).

Notably, Meta’s own advertising standards prohibit adverts promoting tobacco or nicotine products, explicitly including nicotine pouches, which was in force throughout our observation window (23). Pouch adverts identified here therefore point to limited enforcement of this policy.

This study has several limitations. First, reach data from the Meta Ad Library are estimates rather than exact counts, and we could not verify their precision (14). Second, our analysis was limited to Meta platforms (Facebook and Instagram) and does not capture advertising on other platforms such as TikTok, where nicotine pouch promotion has also been documented and tends to have younger user base (7, 24).

Third, we analysed reach at the level of the advert rather than at the level of the individual user, meaning we cannot assess cumulative exposure or frequency of exposure for specific demographic groups, and Meta does not report reach for other gender identities. Fourth, we cannot distinguish the specific mechanisms by which the algorithm produces male-skewed delivery (11). Fifth, demographic information in the Meta Ad Library is based on the age and gender recorded on users’ accounts, which may not correspond to users’ actual characteristics, and some reach attributed to adults may include younger adolescents. Finally, our comparison sample of general adverts provides a useful benchmark but may not fully represent the diversity of advertising on Meta platforms, because random sampling of all adverts was not possible and a small number of high-volume advertisers accounted for much of the sample.

In conclusion, this study provides novel evidence that the reach of nicotine pouch advertising on social media was substantially skewed towards young men, even when adverts were not explicitly targeted at this group. These findings suggest that regulation focused solely on advertiser-set targeting settings may not adequately capture how advertising exposure is distributed in practice.

## Data Availability

All data produced are available online at https://www.facebook.com/ads/library/api/

## References

1. Grandolfo E, Ogden H, Fearon IM, Malt L, Stevenson M, Weaver S, Nahde T. Tobacco-Free Nicotine Pouches and Their Potential Contribution to Tobacco Harm Reduction: A Scoping Review. Cureus. 2024;16(2):e54228.

2. Travis N, Warner KE, Goniewicz ML, Oh H, Ranganathan R, Meza R, et al. The Potential Impact of Oral Nicotine Pouches on Public Health: A Scoping Review. Nicotine Tob Res. 2025;27(4):598–610.

3. Sun H, Tattan-Birch H, Oldham M, Cox S, Jackson SE. Oral Nicotine Pouch Use Among U.S. Middle and High School Students, 2021-2023. medRxiv. 2026:2026.01.28.26345040.

4. Tattan-Birch H, Jackson SE, Shahab L, Buss V, Sun T, Read D, et al. Oral nicotine pouch use in Great Britain: a repeat cross-sectional study, 2020-25. Lancet Public Health. 2026;11(1):e26–e34.

5. Taylor E, Ebdon M, Nottage M, Simonavicius E, Brose L, McNeill A, et al. Investigating the Appeal of Nicotine Pouch Packaging, Flavour, and Nicotine Descriptors Among Adults in the UK: An Online Experiment. Nicotine Tob Res. 2026;28(2):251–9.

6. Moodie C, Alexandrou G, Siddiqi K. Chasing a buzz: developments in the nicotine pouch market in the UK. Tob Control. 2025;34(5):708–9.

7. Sun T, Tattan-Birch H. Sports, Gigs, and TikToks: Multichannel Advertising of Oral Nicotine Pouches. Nicotine Tob Res. 2026;28(2):304–8.

8. Leadbitter G. Nicotine: Advertising - Written question 122419. UK Parliament Written Questions and Answers. 2026.

9. Advertising Standards A. Pouches, crushballs and beyond: an update on the ASA’s recent work on e-cigarettes and related products. CAP News. 2022.

10. Tobacco and Vapes Act 2026, (2026).

11. Ali M, Sapiezynski P, Bogen M, Korolova A, Mislove A, Rieke A. Discrimination through Optimization: How Facebook’s Ad Delivery Can Lead to Biased Outcomes. Proceedings of the ACM on Human-Computer Interaction. 2019;3:1–30.

12. Xiao LY, Deery C, Petrovskaya E, Park S, Newall P. Widespread illegal video game advertising in the UK and South Korea: Many adverts not disclosing loot box presence found using Meta’s ad repository. Journal of Behavioral Addictions. 2025;14(2):714–23.

13. Xiao LY, Deery C. A significant minority of Dutch gambling adverts on social media illegally targeted and likely reached under-24s: Assessing compliance using Meta’s advertising repository mandated by the EU Digital Services Act. J Behav Addict. 2026;15(2):833–45.

14. Meta. Meta Ad Library API 2026 [Available from: https://www.facebook.com/ads/library/api/.

15. Meta. Continuing to create age-appropriate ad experiences for teens. 2023.

16. Haldane BJBS. THE ESTIMATION AND SIGNIFICANCE OF THE LOGARITHM OF A RATIO OF FREQUENCIES. Annals of Human Genetics. 1956;20(4):309–11.

17. Efron B, Tibshirani RJ. An Introduction to the Bootstrap. 1st ed: Chapman and Hall/CRC; 1994 1994.

18. Votta F, Dobber T, Guinaudeau B, Helberger N, de Vreese C. The Cost of Reach: Testing the Role of Ad Delivery Algorithms in Online Political Campaigns. Political Communication. 2024;42(3):476–508.

19. Petrovskaya E, Xiao LY, Khoo N, Leahy D, Roberts A. Gambling adverts on social media reach 2.3 times more men than women: Using the Meta Ad library to assess gambling advertising in Ireland. Journal of Behavioral Addictions. 2026:2006.25.00484.

20. Parliament U. Tobacco and Vapes Bill London: UK Parliament; 2025 [Available from: https://bills.parliament.uk/bills/3879.

21. Imana B, Shen Z, Heidemann J, Korolova A. External Evaluation of Discrimination Mitigation Efforts in Meta’s Ad Delivery. Proceedings of the 2025 ACM Conference on Fairness, Accountability, and Transparency: Association for Computing Machinery; 2025. p. 2616–29.

22. Duren M, Atella L, Welding K, Kennedy RD. Nicotine pouches: a summary of regulatory approaches across 67 countries. Tobacco Control. 2024;33(e1):e32.

23. Meta. Advertising Standards: Tobacco and Related Products. Meta Transparency Centre. 2024.

24. Zenone M, Harries B, Hartwell G. The Promotion of Oral Nicotine Pouches for Non-Smoking Cessation Purposes on TikTok. Nicotine & Tobacco Research. 2026;28(2):282–6.

