## Supplementary material for "Nicotine pouch adverts reach 12 times more young men than women: targeting and reach on Meta social media platforms in the UK"

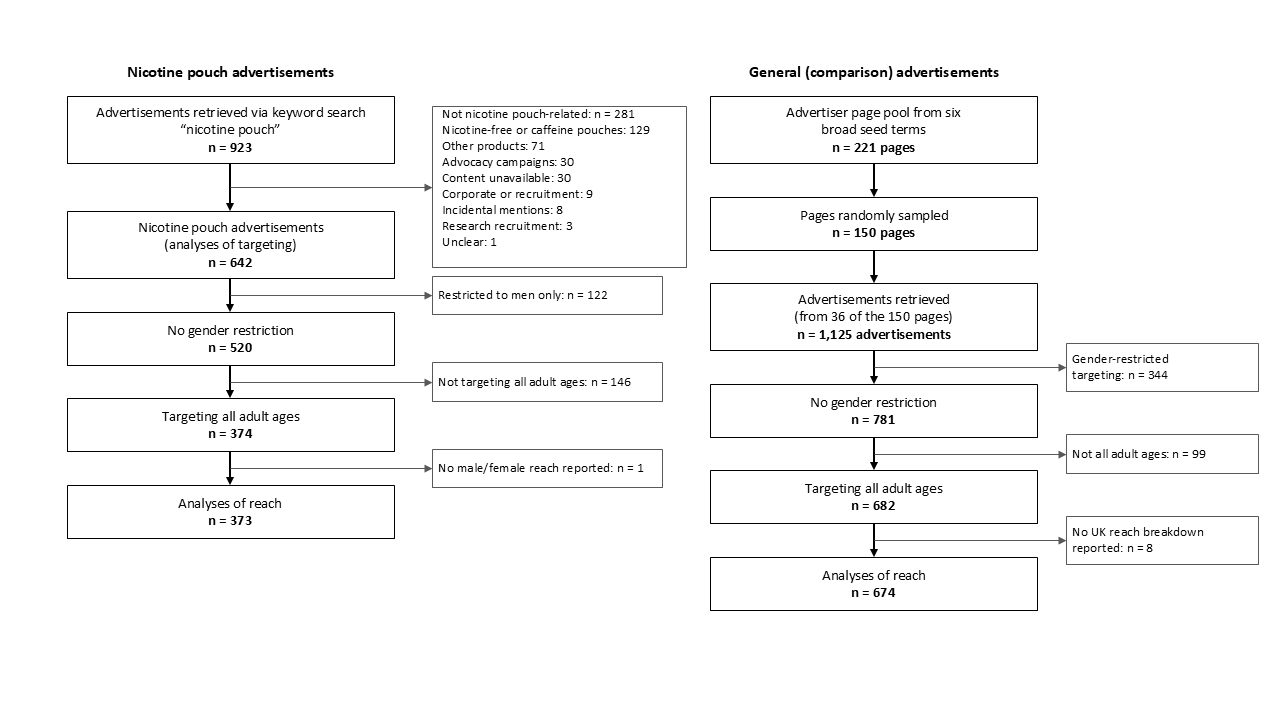

**Supplementary Figure 1.** Flow chart of selecting the nicotine pouch and general advertisements

**Supplementary Table 1.** Number of advertisers and concentration of adverts by advertiser, for nicotine pouch and general advert samples

| **Sample** | **Adverts (n)** | **Advertisers (n)** | **Largest advertiser (%)** | **Five largest advertisers (%)** |
| --- | --- | --- | --- | --- |
| Nicotine pouch adverts, full sample | 642 | 70 | 12.3 | 43.6 |
| Nicotine pouch adverts, reach analysis sample | 373 | 49 | 12.3 | 45.8 |
| General adverts, full sample | 1,125 | 36 | 6.7 | 33.3 |
| General adverts, reach analysis sample | 674 | 22 | 11.1 | 54.2 |

**Supplementary Table 2.** Composition of the general advert comparison sample by advertiser category

| **Advertiser category** | **Advertisers, full sample (n)** | **Adverts, full sample, n (%)** | **Advertisers, reach analysis sample (n)** | **Adverts, reach analysis sample, n (%)** |
| --- | --- | --- | --- | --- |
| Retail and consumer goods | 10 | 424 (37.7) | 10 | 402 (59.6) |
| Fiction and reading apps | 7 | 325 (28.9) | 1 | 1 (0.1) |
| Health and wellness | 8 | 263 (23.4) | 6 | 235 (34.9) |
| Entertainment, media and sport | 5 | 66 (5.9) | 2 | 3 (0.4) |
| Business and consumer services | 4 | 37 (3.3) | 2 | 27 (4.0) |
| Food and hospitality | 1 | 6 (0.5) | 1 | 6 (0.9) |
| Charity | 1 | 4 (0.4) | 0 | 0 (0.0) |
| Total | 36 | 1,125 (100.0) | 22 | 674 (100.0) |
| Note: Advertisers were identified by the Facebook page that ran each advert and categorised by one author (HS) on the basis of the page name and the content of its adverts as displayed in the Meta Ad Library. The full sample comprises all general adverts retrieved, and the reach analysis sample comprises those with no advertiser-set gender restriction that targeted all adult ages and for which estimated reach was reported for men and women in the UK.  Adverts run by advertisers in the fiction and reading apps category were almost all restricted by the advertiser to women only (324 of 325) and were therefore excluded from the reach analysis sample by the same criteria applied to nicotine pouch adverts. | | | | |

**Supplementary Table 3. Male-to-female reach ratios (RR) by age group for nicotine pouch adverts versus general adverts on Meta platforms**

| **Age group** | **Male vs female RR** | | **RRR** |
| --- | --- | --- | --- |
|  | **Pouch adverts**  **(95% CI)** | **General adverts**  **(95% CI)** | **Pouch vs general (95% CI)** |
| 18–24 | 12.51 (10.93, 14.36) | 0.81 (0.70, 0.95) | 15.38 (12.31, 18.82) |
| 25–34 | 11.80 (10.21, 13.54) | 0.69 (0.59, 0.79) | 17.22 (13.99, 21.16) |
| 35–44 | 8.05 (7.10, 9.20) | 0.59 (0.51, 0.70) | 13.56 (10.98, 16.57) |
| 45–54 | 4.88 (4.25, 5.65) | 0.55 (0.47, 0.65) | 8.83 (7.08, 10.90) |
| 55–64 | 3.12 (2.80, 3.52) | 0.52 (0.45, 0.61) | 6.03 (4.93, 7.26) |
| 65+ | 3.05 (2.66, 3.50) | 0.63 (0.54, 0.72) | 4.86 (3.98, 5.93) |

**Note:** RRs compare reach of adverts to men versus women within each age group (values >1 indicate greater reach to men). RRRs compare this difference for nicotine pouch adverts versus general adverts. Analyses were restricted to adverts with no advertiser-set gender restriction that targeted all adult ages. Estimates are geometric means across adverts with 95% confidence intervals (CIs) from bootstrap resampling. N for pouch adverts = 373, and N for general adverts = 674.

**Supplementary Table 4.** Male-to-female reach ratios (RR) by age group, based on total estimated reach summed across nicotine pouch adverts (n = 373)

| **Age group** | **Female reach** | **Male reach** | **Male/Female RR** |
| --- | --- | --- | --- |
| 18–24 | 362,491 | 1,824,825 | 5.03 |
| 25–34 | 741,015 | 2,504,372 | 3.38 |
| 35–44 | 587,410 | 1,308,218 | 2.23 |
| 45–54 | 425,908 | 803,669 | 1.89 |
| 55–64 | 489,749 | 675,617 | 1.38 |
| 65+ | 509,908 | 492,427 | 0.97 |
| Note: Male-to-female RRs were calculated by dividing total male reach by total female reach across all adverts within each age group, giving greater weight to adverts with higher reach. Analyses were restricted to adverts with no advertiser-set gender restriction that targeted all adult ages. Adverts included were those shown in the UK on Meta platforms in the 12 months preceding 8 December 2025. | | | |

**Supplementary Table 5.** Male-to-female ratios of reach ratios (RRR) with confidence intervals accounting for clustering of adverts within advertisers

| **Age group** | **RRR** | **Advert-level 95% CIᵃ** | **Advertiser-level 95% CIᵃ** |
| --- | --- | --- | --- |
| 18–24 | 15.38 | (12.31, 18.82) | (5.24, 43.43) |
| 25–34 | 17.22 | (13.99, 21.16) | (6.31, 50.31) |
| 35–44 | 13.56 | (10.98, 16.57) | (4.64, 44.18) |
| 45–54 | 8.83 | (7.08, 10.90) | (2.71, 28.67) |
| 55–64 | 6.03 | (4.93, 7.26) | (2.01, 17.88) |
| 65+ | 4.86 | (3.98, 5.93) | (1.70, 12.70) |
| ᵃ Confidence intervals were derived from 1,000 bootstrap resamples.  RRR compares the male-to-female reach ratio for nicotine pouch adverts with that for general adverts. Point estimates are identical under both bootstrap schemes while the resampling unit differs (individual adverts vs advertiser pages). | | | |

**Supplementary Table 6.** Age-group ratios of reach ratios (RRR) accounting for clustering of adverts within advertisers

| **Age group** | **RRR** | **Advert-level 95% CIᵃ** | **Advertiser-level 95% CIᵃ** |
| --- | --- | --- | --- |
| 18–24 | 7.91 | (6.56, 9.50) | (3.96, 15.71) |
| 25–34 | 2.46 | (2.20, 2.75) | (1.38, 4.18) |
| 35–44 | Ref | Ref | Ref |
| 45–54 | 0.67 | (0.59, 0.75) | (0.39, 1.25) |
| 55–64 | 0.46 | (0.38, 0.56) | (0.18, 1.18) |
| 65+ | 0.66 | (0.53, 0.82) | (0.18, 2.05) |
| ᵃ Confidence intervals were derived from 1,000 bootstrap resamples.  RRR compares the reach ratio for each age group relative to 35–44-year-olds for nicotine pouch adverts with the corresponding ratio for general adverts. Point estimates are identical under both bootstrap schemes while the resampling unit differs (individual adverts vs advertiser pages). | | | |
